# Assessing the Global Tendency of COVID-19 Outbreak

**DOI:** 10.1101/2020.03.18.20038224

**Authors:** Qinghe Liu, Zhicheng Liu, Junkai Zhu, Yuhao Zhu, Deqiang Li, Zefei Gao, Liuling Zhou, Yuanbo Tang, Xiang Zhang, Junyan Yang, Qiao Wang

## Abstract

COVID-19 is now widely spreading around the world as a global pandemic. In this report, we estimated the global tendency of COVID-19 and analyzed the associated global epidemic risk, given that the status quo is continued without further measures being taken.

Based on official data of confirmed and recovered cases until May 21, 2020, the results showed that the global *R*_0_, excluding China, was estimated to be 2.76 (95% CI: 2.57 – 2.95). The United States, Germany, Italy, and Spain have peak values over 100,000. Using dynamical model and cluster analysis, we partition the globe into four regional epicenters of the outbreak: Southeast Asia extending southward to Oceania, the Middle East, Western Europe, and North America. Among them, Western Europe would become the major center of the outbreak. The peak values in Germany, Italy, and Spain were estimated to be 228,000, 291,000, and 298,000, respectively. Based on the current control measures by May 21, 2020, the peak value in the United States will reach 2,114,000. The cumulative number of 51 mainly researched countries’ patients might finally attain 6,542,000 (95% CI: 4,772,000 – 40,735,000). We also estimated the diagnosis rate, recovery rate, and infection degree of each country or region, and used clustering algorithm to retrieve countries or regions with similar epidemic characteristics. Several suggestions have been proposed for countries or regions in different clusters.

## 1. INTRODUCTION

On Dec. 8, 2019, the first case of the novel coronavirus pneumonia (NCP) was confirmed in Wuhan, Hubei Province, China, caused by a new type of coronavirus named ‘2019 new coronavirus [1] (SARS-CoV-19)’ [2]. The disease spread across China during the traditional Chinese Spring Festival. At 24:00 on March 14, China had passed the peak of the epidemic, and started to recover gradually, with a total of 80,844 confirmed cases reported [3]. However, new cases of disease began to appear in other parts of the world and increased rapidly. Because of the rapid spread of the disease, the World Heath Organization (WHO) has assessed COVID-19 as a pandemic [4], which is the first pandemic caused by a coronavirus. As of March 14, 2020, the cases of COVID-19 have been reported in 135 countries and regions worldwide, with a total of 142,539 confirmed cases (61,518 outside of China) and 5,393 deaths (2,199 outside of China) [5]. Among them, a total of 17,660 cases were confirmed in Italy, 11,364 in Iran, 8,086 in the Republic of Korea, 1,678 in the United States. Furthermore, Europe became the epicenter of the pandemic, with more reported cases and deaths than the rest of the world combined, apart from China [6].

With the outbreak of COVID-19, countries or regions have been taking different measures to cope with the spread of the pandemic, but the number of infected people is still increasing. Meanwhile, the spread of the pandemic has also caused a huge impact on the trade flows and economic affairs of the world. These situations raised many urgent problems. How will this epidemics spread in countries or regions around the world? When will the spread of epidemic arrive at the peak or turn to stabilize? How many people will be at risk of infection? Therefore, it is of great significance to analyze the trend of COVID-19 and predict the arrival of peaks for the prevention and control of the pandemic all over the world.

Based on global data [7], we perform an analysis of epidemic status in 51 countries, which had the number of confirmed cases over 100 in Mar. 27, 2020. Section 2 of this paper briefly discusses the most up-to-date literature related to the COVID-19 epidemic status. Section 3 describes the transmission dynamics model in detail. Section 4 presents the simulation and analysis of the estimated trend. Section 5 analyzes the four epidemic transmission clusters with different characteristics and give error analysis for confirming the rationality of the results. Finally, the conclusion and discussion are respectively presented in Sections 6 and 7.

## 2. RELATED WORK

With the further spread of the epidemic, many scholars analyzed the characteristics of covid-19. On March 1, 2020, Li Y *et al* [8], used the outside-China diagnosis number released by the WHO and built a mathematical model to capture the global trend of epidemics outside China. They found that 34 patients outside China were not found. The worldwide epidemic trend is approximately exponential, and may grow 10 folds every 19 days. Zhuang Z *et al* [9], used a stochastic model to simulate the transmission process of South Korea and Italy under two corresponding assumptions of exponential growth periods. The results indicated that the reproductive number of South Korea and Italy were 2.6 (95% CI: 2.3-2.9) and 3.3 (95% CI: 3.0-3.6), respectively. On March 17, Darwin R *et al* [10], used the progression of the epidemic curve and defined frame work to estimate the instantaneous reproductive number. They divided the epidemic into different serial interval scenarios, and estimated the *R* values, average number of secondary transmissions per patient. The *R* value in Japan, Germany, Spain, Kuwait, and France are over 2; in Italy, Iran, and South Korea are even over 10, however, the R will be low after the social distancing intervention.

The correlation analysis of COVID-19’s various factors is also an important topic. Cody Carroll, Satarupa Bhattacharjee, Yaqing Chen, *et al* [11] established a framework for quantifying and comparing cases and deaths across countries longitudinally. They found that decreased workplace mobility is associated with lower doubling rates with a roughly two week delay, and case fatality rates exhibit a positive feedback pattern. Alessio Notari, Giorgio Torrieri [12] analyzed risk factors correlated with the initial transmission growth rate of the recent COVID-19 pandemic in different countries on May 8, 2020 and found moderate evidence for positive correlation with: CO2 (and SO) emissions (p-value 0.015), type-1 diabetes in children (p-value 0.023), vaccination coverage for Tuberculosis (BCG) (p-value 0.028).

In addition, many scholars have also discussed different models. Andrey Gerasimov, Georgy Lebedev, Mikhail Lebedev and Irina Semenycheva proposed a heterogeneous model [13] on May 7, 2020 to assess the effects of different measures for infection risk control. They found that after this heterogeneity is incorporated in the model, several characteristics of the epidemic are estimated more accurately. For higher accuracy, Sina F. Ardabili, Amir Mosavi, Pedram Ghamisi, *et al* [14] used multi-layered perceptron and adaptive network-based fuzzy inference system to analyze COVID-19, which got better results than traditional methods. On May 1, 2020, Kevin L *et al* [15] proposed a dynamic SEIR epidemiology model with a time-varying reproduction number, which they identify using machine learning and uncertainty quantification. This new dynamic SEIR model provides the flexibility to simulate various outbreak control and exit strategies to inform political decision making and identify safe solutions in the benefit of global health.

The prediction of pandemic was also a hot spot. Zhang Z *et al* [16] estimated that the peaks of infectious cases in South Korea, Italy, and Iran are expected to occur at the end of March, and the percentages of population infections will less than 0.01%, 0.05%, and 0.02%, respectively. On March 10, 2020, Zhang Z *et al* [17] estimated that the inflection point arrival time will be March 6-12 for South Korea, March 10-24 for Italy and March 10-24 for Iran, and the cumulative number of cases will reach 20k in South Korea, 209k in Italy and 226k in Iran, respectively. Li L *et al* [18] thought that the epidemic in South Korea will be basically under control at the end of March, and the inflection day is January 7, before the control the reproductive number is 4.2, and 0.1 after the control. Epidemic size in Italy and Iran will reach 200,000 and 20,000, respectively at the end of March. On May 8, 2020, Athanasios S *et al* [19], found that the rate of deaths reaches a maximum, their models provide estimates for the time that a plateau will be reached signifying that the epidemic is approaching its end, as well as for the cumulative number of deaths at that time. The plateau is defined to occur when the rate of deaths is 5% of the maximum rate.

In our previous work [20], we predicted the time and value of the inflection point and peak point in both Hubei and outside Hubei of China. Inspired by it, we conducted a worldwide prediction of the value of the peak point and analyzed the main characteristics and appearances of COVID-19 in the world. The predicted results are based on the current medical situation, if there is specific medicine for COVID-19 in the future, the time and value of peak point of all countries will be advanced and shrinking compared with non-medicine situations.

## 3. COVID-19 TRANSMISSION MODEL: TRANSMISSION DYNAMICS

### 3.1. Transmission Dynamics Model

We constructed a transmission dynamics model to infer the epidemiological characteristics and the peak size and trend of COVID-19 based on the existing infectious data and recovered data about various countries or regions. The population of this paper are divided into four main categories based on the SEIR model, i.e. *S, E, I*, and *R* referring to [21]: the Susceptibles, Exposed, Infectious, and Recovered, respectively [22][23]. In the study of China’s epidemic situation, many factors such as Spring Festival and control policies and so on are taken into account, but in contrast, this paper will analyze the heterogeneity of each country or region on the basis of the common spread of infectious diseases. Although these countries have different policies and customs, their basic spread pattern of the epidemic still meets the principle of dynamics [24]. S, the susceptibles, is equal to the total population (N in equation (3.1)) in the research of COVID-19 due to the general susceptibility of the population. The main assumptions of this model are as follows:

1. Because the difference in infectivity between the exposed and the infectious population is unknown, both groups of *E* and *I* are set to follow the same coefficient *β* to represent the average infection level of COVID-19.
2. In the predicted time scale, the existing influences of policies or culture of the target object will be unchanged.
3. During the transmission of virus, the probability of infection in contact with each person is equal in the same target group.

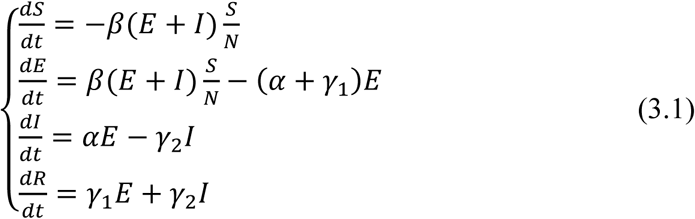

where *t* means time, and *S, E, I, R* represent four different state variables, respectively. Class *S* represents a healthy population. Once transferred to class *E*, it means the population has been infected until transferred to class *R*, recovered. People in class *I* have been diagnosed due to symptom detection. Formula (3.1) can be represented by the state transition diagram in Figure 1. The direction of the arrow in Figure 1 represents the transition relationships between different states with time. All parameters and physical interpretations of the model are shown in Table 1.

**Figure 1.**
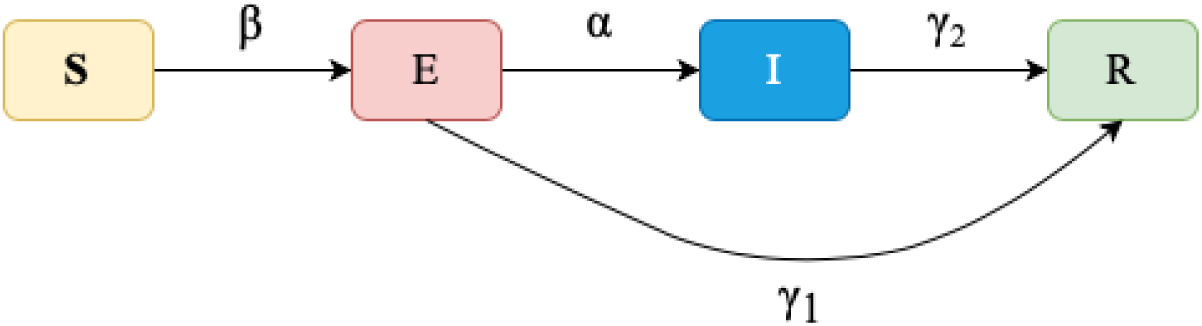
SEIR system state transition diagram.

**Table 1.** List of Symbols.

|  |  |  |
| --- | --- | --- |
| *Constant* | $N$ | Total population of object of study. |
| *State variable* | $S$ | Number of susceptible people, who haven't been infected. |
| | $E$ | Number of exposed people, who are in latent period. |
| | $I$ | Number of infectious people. |
| | $R$ | Number of recovered people. |
| *Transition variables* | $\alpha$ | Rate of those exposed and being infectious. |
| | $\beta$ | Rate of those susceptible and being infected. |
| | $\gamma_1$ | Transmission rate to the recovered from the exposed. |
| | $\gamma_2$ | Transmission rate to the recovered from the infectious. |

### 3.2 Practical Consideration

We divided the spread of the COVID-19 into three phases according to the evolution of the size of the confirmed cases, and the schematic diagram of each stage for the number of patients is shown in Figure 2.

**Figure 2.**
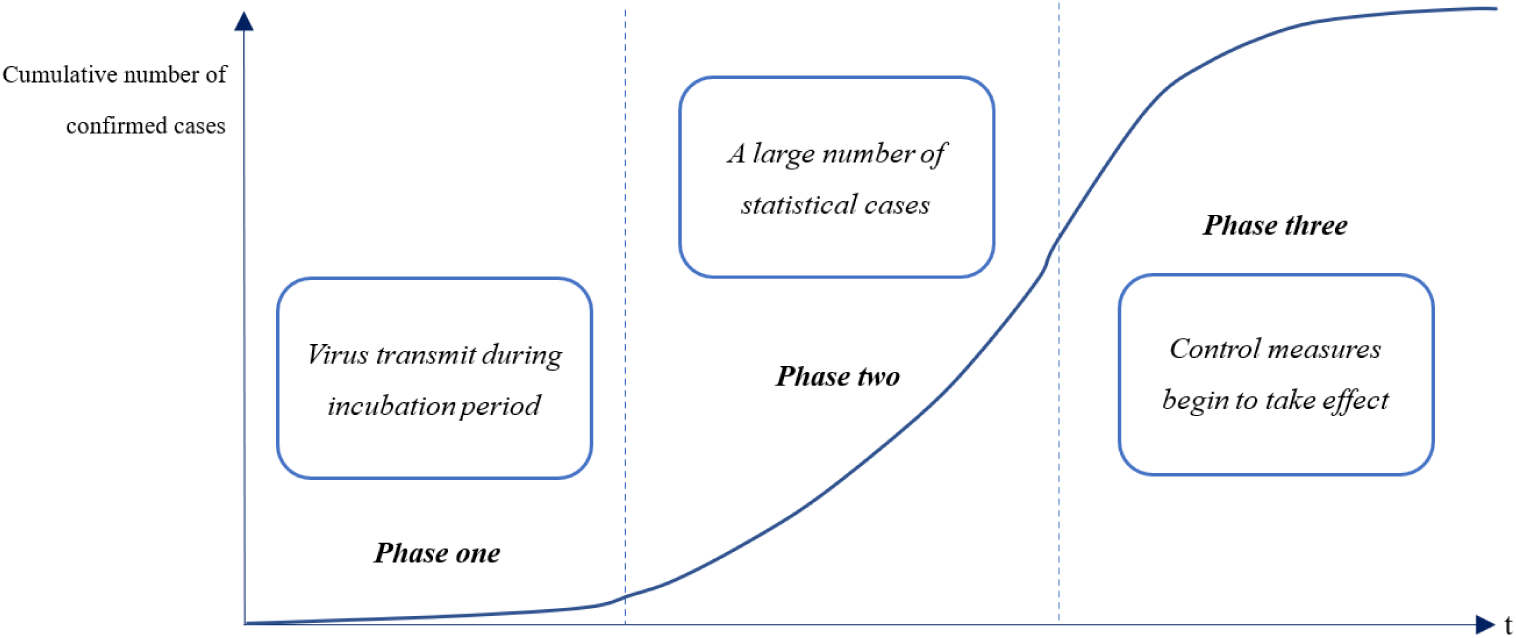
Phase diagram of confirmed cases changing with time in a certain region.

Phase one: the time range of this phase starts from the beginning of COVID-19 transmission and ends when the number of patients begins to increase significantly. COVID-19 spreads during the incubation period at this phase, but the confirmed cases officially counted remain at a low level, and the public’s awareness of defense is light.

Phase two: the main feature is that a large number of patients have been diagnosed, and the statistical data of confirmed cases increase significantly. But there still has no policy intervention and large-scale quarantine measures.

Phase three: As the intensity of policy intervention increases, the spread of COVID-19 is effectively suppressed, and growth rate of confirmed cases is gradually controlled until the end of the epidemic. The main feature of this phase is strong human intervention.

It is crucial to get as much information as possible from the data in three stages. Because this paper aims at the analysis of the peak size and its arrival time in each country or region, two latter transmission phases will be our focus in this paper.

In phase two, SARS-CoV-2 spread rapidly. The infection rate remains at a high level, and the recovery level is low because the government had not carried out obviously effective policy control. From phase three, the infection rate decreases and the recovery rate begins to increase. Through the study of reported data of COVID-19 in China, we used the power law to simulate the trend of infection rate and recovery rate in phase three, as shown in equation (3.2). Since the coefficient c plays a role in inhibiting the growth of the number of confirmed cases, we define c as the incremental inhibition ratio.

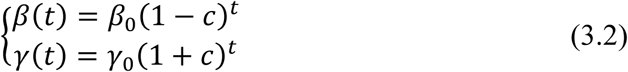

where *β_0_* and *γ*_0_ are the initial values of β and γ, and t is time from the beginning of phase three.

### 3.3. Estimation of *R*_0_

The basic reproduction number, *R*_0_, refers to the average value of how many people an infected person can transmit the virus through natural transmission without external intervention [25]. From the second generator approach in [26], the equation (3.3) is derived as follows:

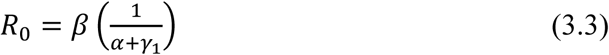

where *α*, *β*, and *γ*_1_ are the transfer variables of the SEIR model. (1*/α + γ*_1_) represents duration of virus transmission. Considering that the calculation of *R*_0_ is in the case of natural transmission, so we use the parameters of the starting time of epidemic spread in each country or region to estimate *R*_0_ to avoid the impact of human intervention.

### 3.4. Data Source

In this study, we used open dataset from Johns Hopkins University (https://github.com/CSSEGISandData/COVID-19). The details of the data source are shown in Appendix 1.

*Johns Hopkins University*. Johns Hopkins University shared their data on GitHub for academic and scientific research. We have compiled their open data into a chronology of confirmed data for prediction and visualization use.

## 4. MAIN RESULTS OF THE SEIR MODEL

### 4.1. Estimate of *R*_0_

While estimating the *R*_0_ of the epidemic globally, we ignore the countries whose ultimate cumulative number of confirmed cases are less than 5000 by May 21, 2020 and focus on the remaining 51 countries (please refer to appendix). Parameters were estimated by minimizing the least square loss function of the estimated number of confirmed cases and the observed number of confirmed cases. The average basic reproduction number of these countries was 2.76 (95% CI: 2.57 – 2.95). As we can notice, COVID-19 is lashing a great part of the world, and the sufferings are not likely to end soon. By estimating the average value of *R*_0_, we obtain the objective information of current situations as well as the importance of epidemic prevention.

Among these 51 mainly researched countries, the *R*_0_ of Switzerland, Austria, China and Brazil are higher than 3.5, suggesting the great potential of a destructive outburst. France is one of these countries with a basic reproduction number of 3.61, much higher than the global average(See Appendix 2 for more details). The number of the United States is 2.59, which is slightly below the global average, revealing the urgency of the current outbreak in the United States. The lowest value of *R*_0_ is 1.77 in Korea, still greater than 1.

**Figure 3.**
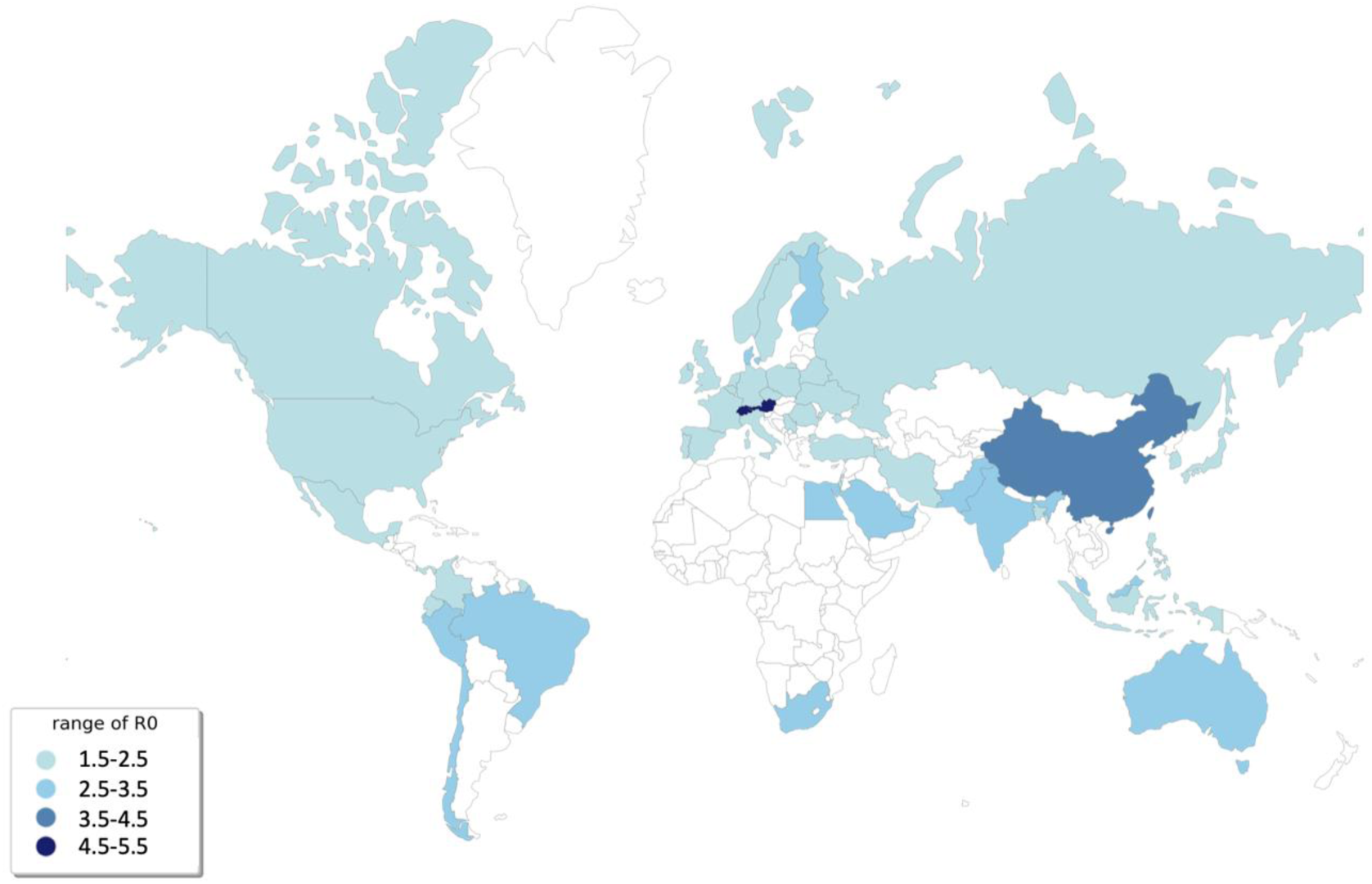
*R*_0_ in 51 countries or regions over the world, which has the number of confirmed cases exceeding 100 by May 21, 2020. Blue means the higher the value of *R*_0_, while white means the lower the the value of *R*_0_.

### 4.2. Estimate of the Peak of Confirmed Cases in Country-Level

The peak value is defined as the cumulative number of confirmed cases. After this peak point, the temporary number of cases gradually decrease or slightly fluctuate, and the epidemic subsides [20]. The estimated trends of 51 countries are all listed in Appendix 2 for details.

Thirteen countries will have a peak value over 100,000, including the United States, Russian Federation, Brazil, the United Kingdom, Spain, Italy, France, Germany, Turkey, Iran, India, Peru and Canada. And 34 of the 51 countries including Belgium, Saudi Arabia, Mexico will have peak values between 10,000 and 100,000.

We also compared peak arrival time in countries whose peak values are higher than 10,000, and find that most of the countries will reach their peak value in June. The United States is the country with the highest exposure to the COVID-19 risk of the coronavirus. According to our estimation, the peak number of real-time confirmed cases of the United States is predicted to reach 2,114,000 after June 18, 2020, provided the present coping strategy is not changed. It is the only country in our prediction that the number of confirmed cases will exceed 2,000,000.

**Figure 4.**
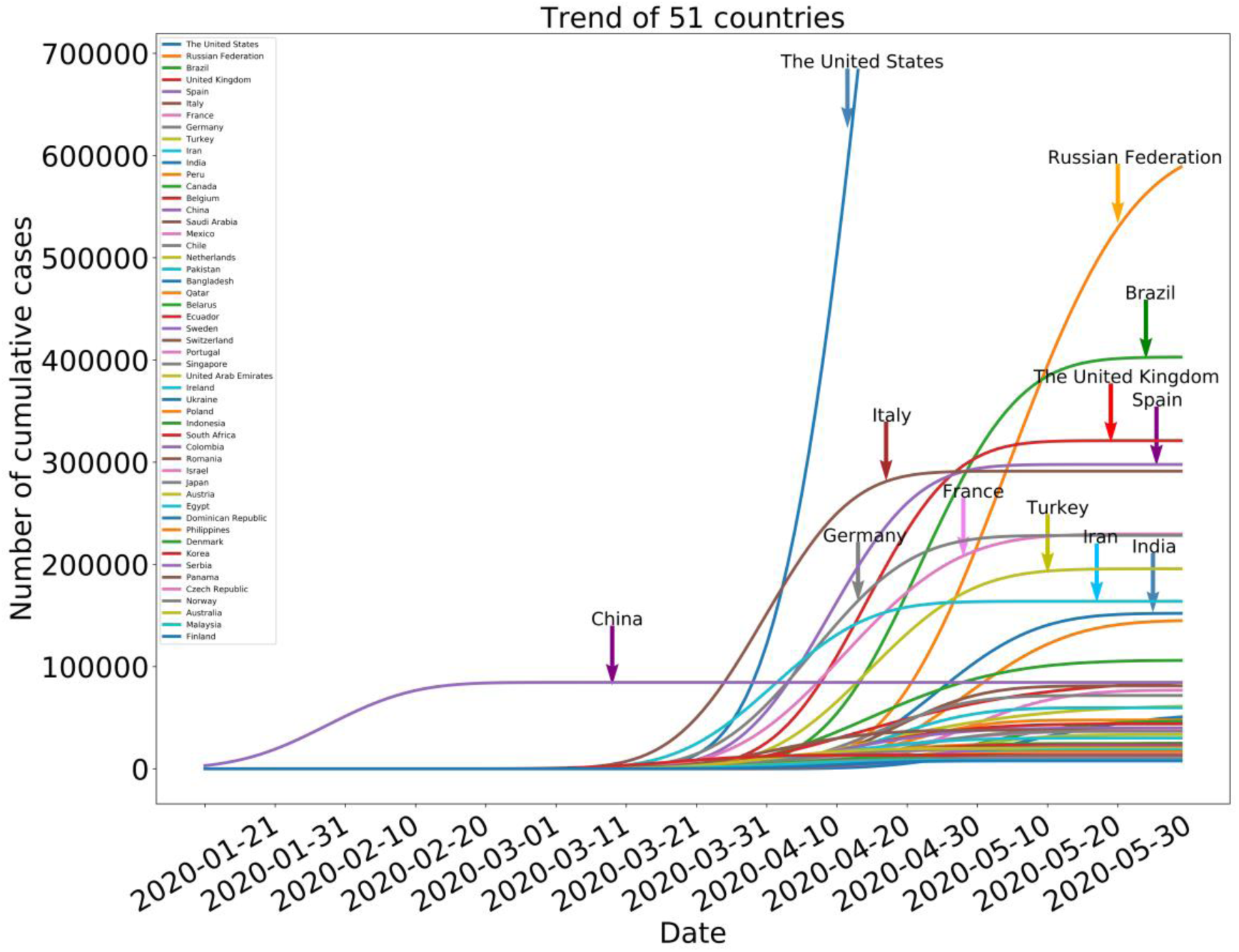
Simulated trends of the cumulative number of confirmed cases in 51 countries or regions, which has the number of confirmed cases exceeding 5000 by May 21, 2020. The horizontal axis is the date, and the vertical axis is the number of confirmed cases.

**Table 2.** Sum of peak values of different continents.

| Peak interval | Countries or regions |
| --- | --- |
| >100,000 | United States, Russian Federation, Brazil, United Kingdom, Spain, Italy, France, Germany, Turkey, Iran, India, Peru, Canada |
| (50,000, 100,000] | Belgium, China, Saudi Arabia, Mexico, Chile, Netherlands, Pakistan, Bangladesh |
| (10,000, 50,000] | Qatar, Belarus, Ecuador, Sweden, Switzerland, Portugal, Singapore, United Arab Emirates, Ireland, Ukraine, Poland, Indonesia, South Africa Colombia, Romania, Israel, Japan, Austria, Egypt, Dominican Republic, Philippines, Denmark, Korea, Serbia, Panama, Czech Republic |
| (5,000, 10,000] | Norway, Australia, Malaysia, Finland |

Spain, Italy, and Germany monopolize one category, out of all 51 countries, due to its high risk of coronavirus. The peak number of real-time confirmed cases of Italy, Spain are respectively predicted to reach 291,000, and 298,000. The trend of COVID-19 in Italy is expanding, though efforts had been taken by the government of Italy. The other countries whose peak values are estimated to reach over 50,000, such as the Bangladesh, Pakistan, Netherlands, Chile, etc., are all in severe situation at present.

### 4.3. Geographical Distribution Analysis of the COVID-19

The estimated final epidemic scale is visualized on the world map to show the geographical characteristics of the predicted epidemic peak of confirmed cases. The cumulative number of 51 countries’ patients might finally attain 6,542,000 (95% CI: 4,772,000 – 40,735,000). According to [27], geographic proximity transmission is very prominent in epidemic transmission, which means that the regional transmission is popular. This opinion can also be confirmed on the map shown in Figure 5. The outbreak shows multiple epicenters. It can be found that North Korea is geographically close to South Korea, Japan, and China, which has the worst situation.

**Figure 5.**
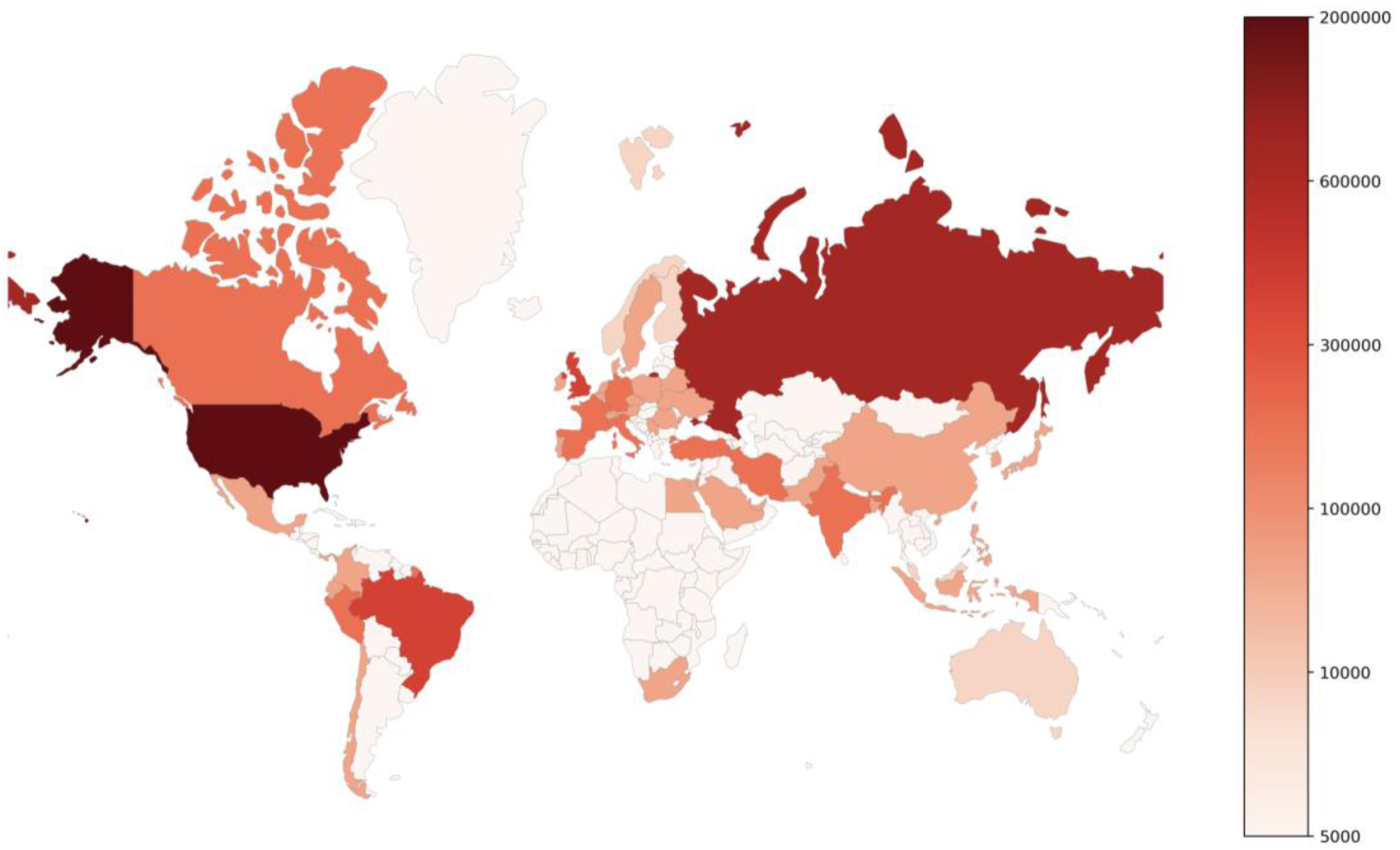
The peak value of confirmed cases in 51 countries or regions over the world, which has the number of confirmed cases exceeding 5000 by May 21, 2020. Red means the higher the diagnosis cases peak, while white means the lower the diagnosis cases peak.

The choropleth map indicates that the spread of COVID-19 will form four major regional clusters. The first epicenter ranges from East Asia to Oceania, including China, South Korea, Japan, Indonesia, the Philippines, Australia, etc. Norway, Australia, Malaysia, Finland will have peak values over 7,000.

The second outbreak epicenter occurs in Western Europe, including Spain, Italy, The United Kingdom, France, Germany, and many other neighboring countries, as well as Turkey. Belgium, Switzerland, Netherlands, Ireland are all expected to have peak values of over 30,000. Considering the number and density of European countries, there would be a serious cross-border phenomenon. Under the current transmission environment and control measures as of May 21, 2020, the estimated number of patients is likely to be at least 2,984,000, with 740 million population in Europe.

The third epicenter was in the Middle East. According to the current trend, the peak number of confirmed cases in Iran will reach to 104,000. According to the current trend, the epidemic situation in Iran has gradually slowed down. In addition, the peak value of Saudi Arabia is estimated to be over 25,000. Estimated by the current situation, the peak of diagnosis in the Middle East will come in early May this year.

The last epicenter occurred in North America. Given that the epidemic has spread in the United States (1,634,000 cases in total, as of May 21, 2020), if the current situation is maintained, the peak number of cases in the United States will reach 2,114,000, which may have a huge impact on American people’s livelihood and social economy. If the United States follows the existing policy as of May. 21, 2020, the peak arrival time of COVID-19 in the United States will be after Jun. 18, 2020. In addition, Canada is expected to have a peak value of 106,000, making it the second most affected country in North America.

### 4.4. Error Analysis of Proposed Model

In this section, the normalized root mean square error (NRMSE) was used to test the accuracy of prediction level of the model. We used the confirmed data as of April 25, 2020 to estimate the data for the following week. The error between predicted value *ŷ* and real value *y* is calculated according to the equation (4.1):

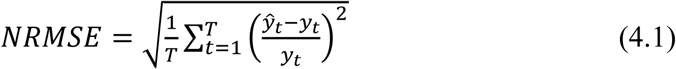

Figure 6 shows the visual display of the error. From a quantitative perspective, the nrmse of the target country or region is 12.85% on average when t = 7. The minimum is 0.42%, while the maximum is 36.97%. And the average error will increase with the increase of prediction time, which is mainly because the longer the time is, the less relevant information is needed for prediction, the more difficult it is, and the higher the cumulative error is.

**Figure 6.**
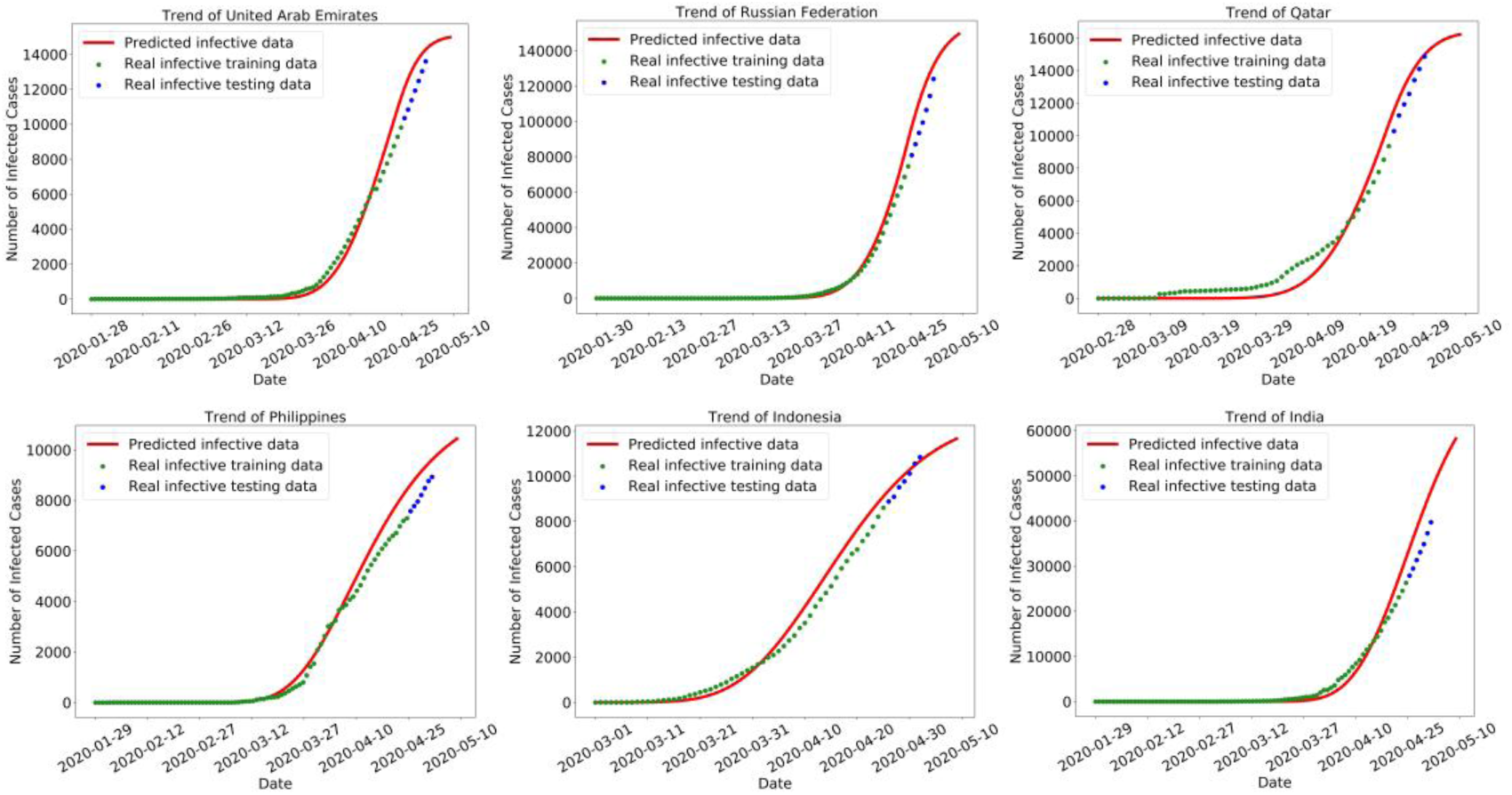
The comparison between the predicted cumulative diagnosis data of 6 countries, including Arabia, Russia and Qatar, and the data reported by the government. The data before April 25, 2020 is used for model training, and the data in the next seven days are predicted.

## 5. CLUSTER ANALYSIS OF EPIDEMIC CHRACTERISTICS

To further analyze the characteristics of epidemic transmission among different countries or regions, we took the five parameters, i.e. *α*, *β, γ*_1_*, γ*_2_*, c*, of the SEIR model obtained in the simulation as feature vectors. Then, the BIRCH (Balanced Iterative Reduction and Clustering Using Hierarchies) algorithm was used to perform clustering based on these feature vectors to retrieve countries or regions with similar epidemic patterns, see Figure 8, and Appendix 3. The average of parameters in each cluster is shown in Table 4. To visualize the proximity relationship of the epidemic characteristics of these countries or regions, t-SNE (t-distributed stochastic neighbor embedding) algorithm is used to reduce the dimension of data to a 2-dimensional plannar, while preserving sample proximity in the 5-dimentional parameter space, see Figure 8.

**Figure 7.**
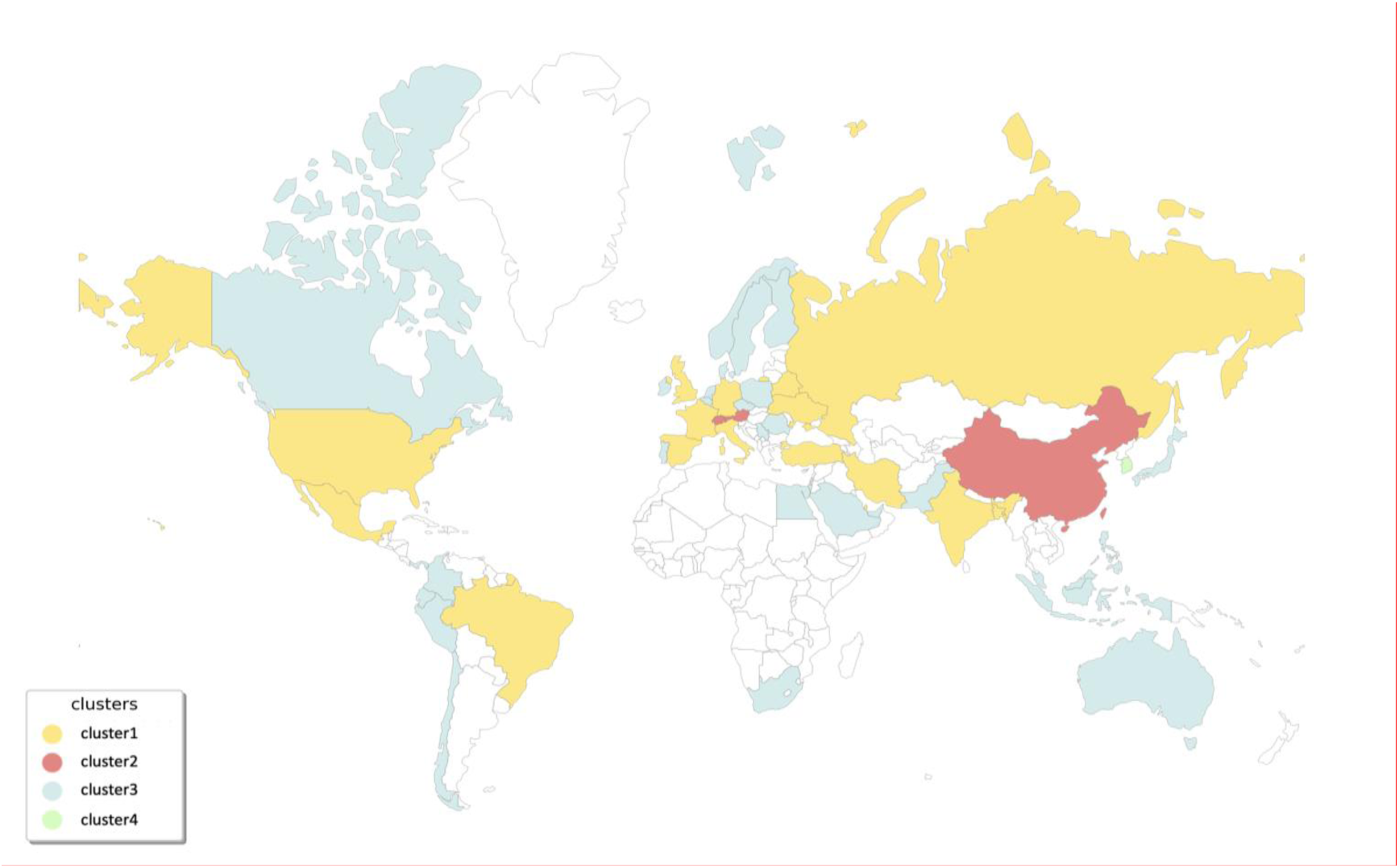
Clustering of the 51 countries or regions with the five SEIR parameters: *α*, *β, γ*_1_*, γ*_2_*, c*,. In this world map, white represents those countries not participating in the clustering.

**Figure 8.**
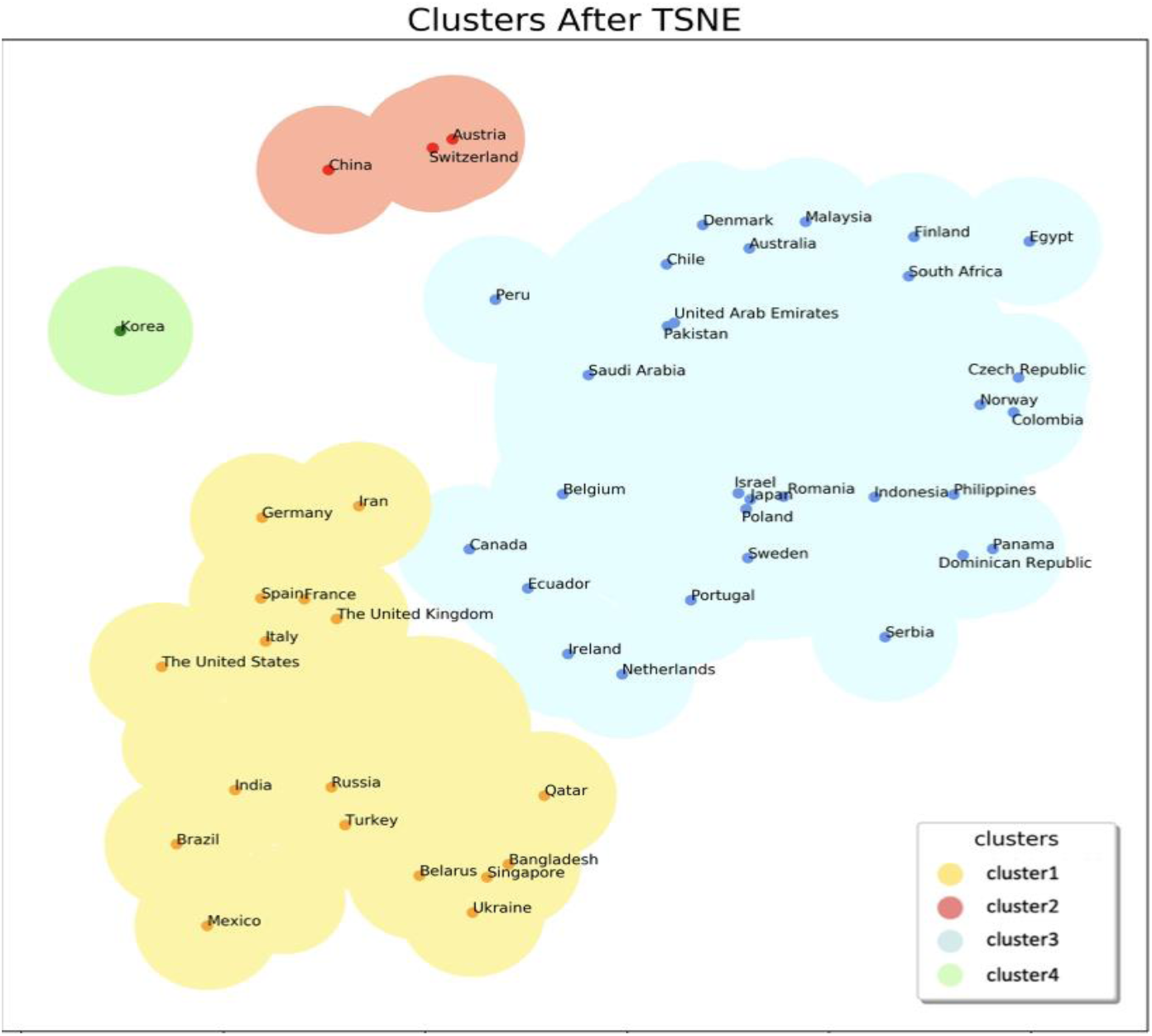
Illustration of SEIR parameter-proximity, i.e. *α*, *β, γ*_1_*, γ*_2_*, c*, of the 51 countries using t-SNE dimension reduction. Owing to the proximity preservation property of t-SNE, the proximity relationship in the 5-dimensional parameter space can be visualized in the above 2-dimensional plannar. Colors indicate the clusters resulted from the clustering algorithm.

**Table 4.** Attributes list of cluster centers.

| Cluster | $\bar{\alpha}$ | $\bar{\beta}$ | $\bar{\gamma}_1$ | $\bar{\gamma}_2$ | $\bar{c}$ | #Countries | Countries or Regions |
| --- | --- | --- | --- | --- | --- | --- | --- |
| One | 0.61 | 1.59 | 0.0065 | 0.0034 | 0.051 | 17 | The United States, Spain, Italy, Germany, etc. |
| Two | 0.30 | 1.52 | 0.0114 | 0.0116 | 0.049 | 3 | Austria, China, Switzerland. |
| Three | 0.54 | 1.43 | 0.0103 | 0.0018 | 0.055 | 30 | Australia, Belgium, Canada, Japan, etc. |
| Four | 0.81 | 1.46 | 0.0098 | 0.0021 | 0.055 | 1 | South Korea. |
| Total | 0.55 | 1.48 | 0.0091 | 0.0033 | 0.053 | 51 | - |

The number of clustering countries or regions indicates the universality or particularity of the epidemic pattern reflected by the cluster. Cluster one is represented by the United States, Italy, Spain, Germany, etc. Their 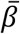 are of the highest level, 1.59. However, 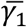 was the lowest at 0.65%, which shows that the epidemic situation in such countries or regions is the most serious, but has not yet got an effective recovery level. It is necessary to remind them to take timely prevention and control measures for the flow of people and invest in medical treatment, otherwise, the epidemic will be likely to further spread.

Cluster two contains only three countries: China, Austria, and Switzerland. Because of the high 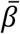, their epidemic spread quickly. But the three countries are excellent at diagnosing and treating patients. 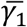 and 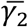 are the highest at 1.14% and 1.16% respectively, which makes their epidemic situation be better suppressed. For example, intervention measures such as traffic blockade and quarantine taken in time at the beginning of the outbreak in China are very effective and have successfully restrained the further growth of confirmed cases. This is also recommended for other countries to learn from.

Cluster three accounts for 59% of the total number of studies, and had the lowest risk among the four clusters. 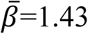 indicates the lowest level of virus transmission, while 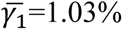 and 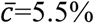 are both higher than the average. Their epidemic spread is very light, and is more easily suppressed. The diagnosis rate, 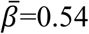, is the most close to the overall average. This includes Australia, Japan, and Belgium, etc.

Cluster four has only one country, South Korea. The diagnosis rate, 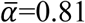, is the highest, which indicates the high demand for diagnosis level. Other parameters were basically very close to the overall average, although the transmission degree is on average, in order to prevent the further spread of COVID-19, the policy of intervention and medical treatment for the confirmed cases still needs to be improved.

It should be noted that clustering is only used to help us find out similar epidemic patterns. It is not that any country or region is fixed as the description of the above four clusters. Figure 8 shows that some countries are at the edge of clustering. The cluster assignments of these countries might be sensitive to the estimated SEIR parameters. We have reason to believe that Peru located at the edge of Cluster Two and Cluster Three also have good epidemic control status. Canada, Ireland, Netherlands, and other countries in cluster three are very close to the first cluster, which indicates the possibility of further outbreaks of COVID-19.

The high risk areas of the outbreak are concentrated in the Americas, Europe, and the Middle East (cluster one). The severity of the epidemic in South Africa is very prominent on the geographical map. On the one hand, we once again call for timely treatment and isolation measures, on the other hand, it is necessary to remind Southern African countries of the prevention of COVID-19 geographic proximity transmission.

**Figure 9.**
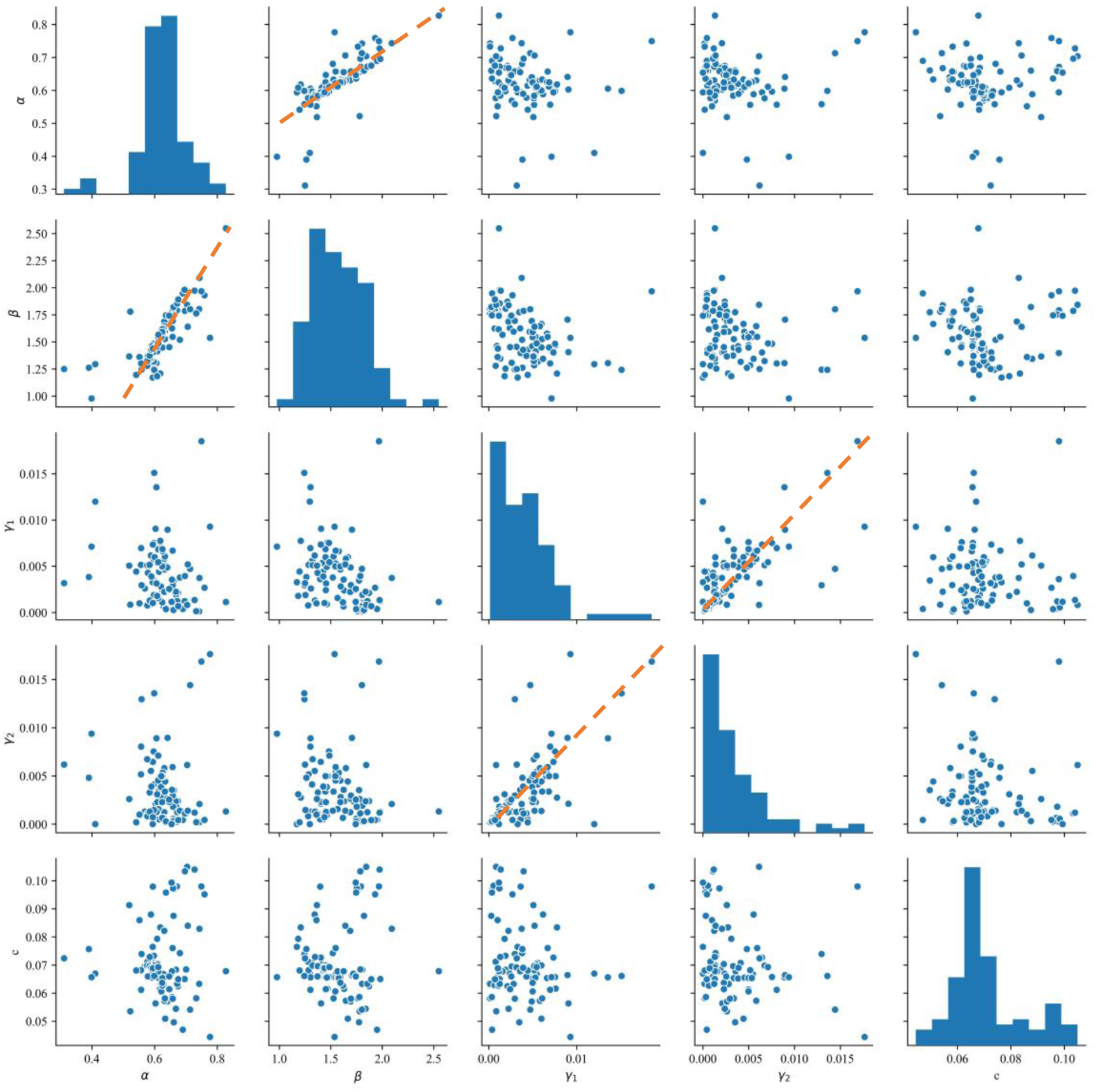
Frequency histograms and scatters of *α*, *β, γ*_1_*, γ*_2_*, c*, which show that there is a linear relationship between *α* and *β*, and there is a linear relationship between *γ*_1_ and *γ*_2_. This shows that countries with rapid COVID-19 transmission are also more efficient in diagnosis, so as to reduce the burden of increasing patients. China not only keep the diagnosis rate at a high level, but also effectively reduce the spreading speed of the epidemic. Most of them have adopted a more timely travel restriction and contact precaution in face with COVID-19.

## 6. CONCLUSION

We performed a simulation analysis of COVID-19 using SEIR model assuming that the current policies by May 21, 2020 remain. From the analysis, we draw the following conclusions:

1. We simulated the transmission of the SARS-CoV-2 in 51 countries. The average basic reproductive number *R*_0_ of these countries was estimated to be 2.76 (95% CI: 2.57 – 2.95).
2. Thirteen countries will have a peak value over 100,000, including the United States, Russian Federation, Brazil, the United Kingdom, Spain, Italy, France, Germany, Turkey, Iran, India, Peru and Canada. And 34 of the 51 countries including Belgium, Saudi Arabia, Mexico will have peak values between 10,000 and 100,000.
3. Western Europe, Southeast Asia to Oceania, the Middle East, and North America will be the four epicenter with the most severe situation. The epidemic situation in North America and Western Europe is estimated to far exceed that in China. In addition, North America will be the worst-hit epicenter.
4. The 51 countries or regions were divided into four clusters by using clustering algorithm based on epidemiology related parameters *α*, *β, γ*_1_*, γ*_2_*, c*. The discovered four clusters represent low risk, medium risk, high risk, and effective control respectively. Based on the similarity of epidemic characteristics, we gave early warnings to many countries, including Canada, Ireland, etc.

## 7. DISCUSSION

In this paper, we mainly estimated and analyzed the transmission trend of COVID-19 based on the daily updated data of confirmed cases and recovered cases. In fact, the trend of COVID-19 is related to multiple factors. China’s prior data were used in the third phase of the epidemic (see Figure 2); however, the testing policies and standards in each country and region are different, which is highly related to the governance system and safety consciousness. In addition, in the second phase of epidemic transmission, community transmission plays an important role. The intensity of people flow directly affects the infection degree of the virus. Some factors, such as subway traffic, catering facilities, and customs or habits of each region remains to be studied.

All the analyses above are based on the assumption that the official statistics of confirmed cases and of recovered cases reflect the real status quo of epedemics. As a matter of fact, there may be deviations in the norms of statistical data in various countries, which may also lead to bias in our results.

## Data Availability

The datasets and code used for the current study are available from the corresponding author on reasonable request. Example code used for the analysis is available on GitHub.

https://github.com/epimath/cm-dag

**Abbreviations**
COVID-19: Coronavirus Disease 2019
SARS-CoV-2: Severe Acute Respiratory Syndrome Coronavirus 2
CI: Confidence Interval
SEIR: Susceptible-Exposed-Infectious-Recovered.

## Declarations

### Ethics approval and consent to participate

Not applicable.

### Consent for publication

Not applicable.

### Availability of data

The datasets used and analyzed during the current study is available from open resources.

### Authors’ Contributions

Conceived and designed the experiments: Qinghe Liu, Junkai Zhu, Junyan Yang, and Qiao Wang.

Performed the mathematical modelling: Qinghe Liu, Junkai Zhu, and Qiao Wang. Analyzed the data: Qinghe Liu, Junkai Zhu, Zhicheng Liu, and Yuhao Zhu.

Collect the data: Zefei Gao, and Deqiang Li.

Performed the computations: Yuanbo Tang, and Xiang Zhang.

Wrote the paper: Junkai Zhu, Qinghe Liu, Deqiang Li, Liuling Zhou, Zefei Gao, Zhicheng Liu, and Yuhao Zhu.

All authors read and approved the final manuscript.

## Acknowledgments

Not applicable.

## Appendix 1 Data Source (All data used in the paper is public.)

https://github.com/CSSEGISandData/COVID-19

## Appendix 2 Simulation Results in 51 Main Countries or Regions

Data used as of May 21, 2020. All countries or regions are in alphabetical order.

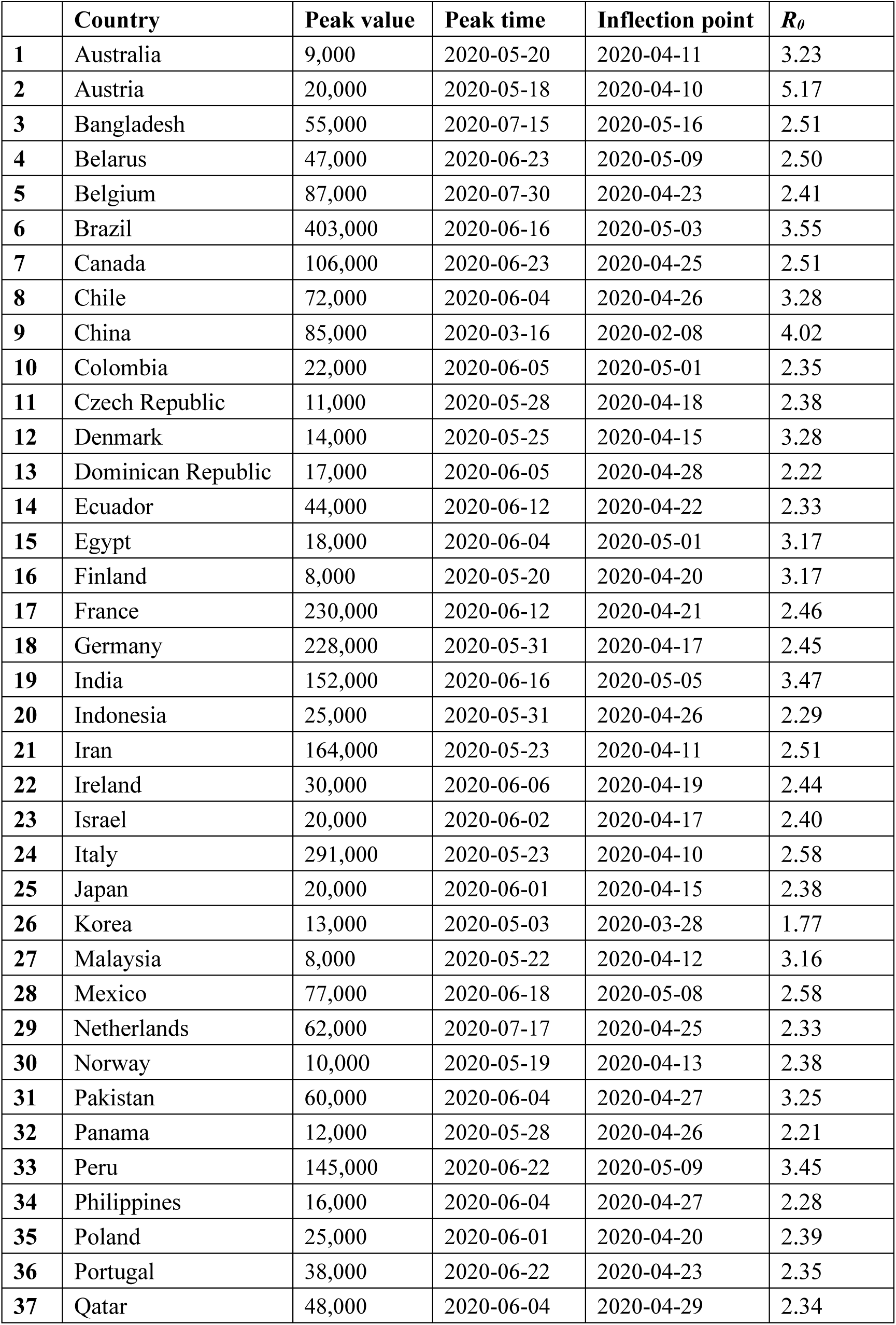
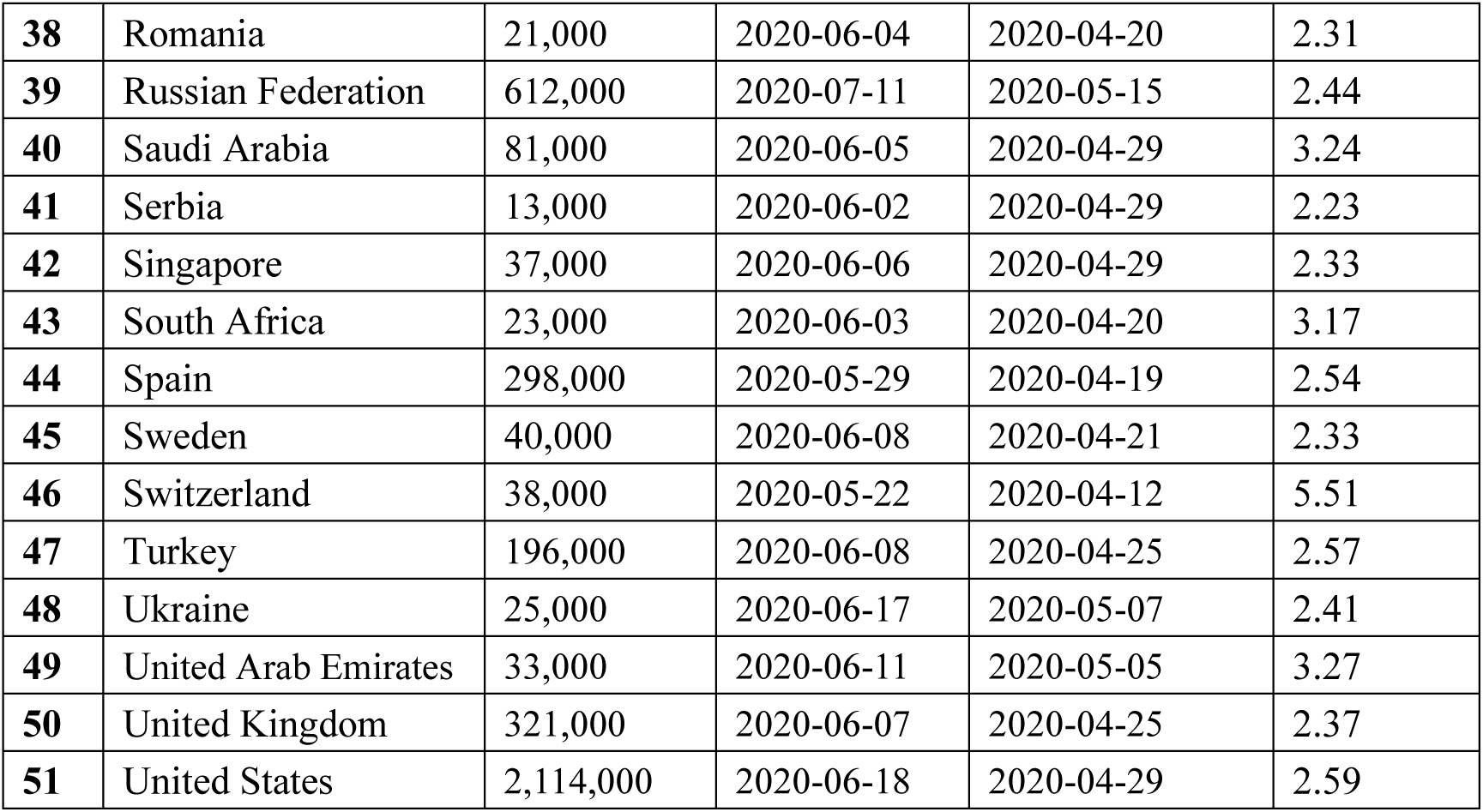

## Appendix 3 Clustering Results and the Estimated SEIR Parameters in 51 Main Countries or Regions

Data used as of May 21, 2020. All countries or regions are in alphabetical order.

Appendix 3 Clustering Results and the Estimated SEIR Parameters in 51 Main Countries or Regions Data used as of May 21, 2020. All countries or regions are in alphabetical order.
| | Country | $\alpha$ | $\beta$ | $\gamma_1$ | $\gamma_2$ | $c$ | Cluster |
| --- | --- | --- | --- | --- | --- | --- | --- |
| 1 | Australia | 0.4361 | 1.4434 | 0.0107 | 0.0052 | 0.0566 | 3 |
| 2 | Austria | 0.2782 | 1.4902 | 0.0100 | 0.0112 | 0.0516 | 2 |
| 3 | Bangladesh | 0.6324 | 1.6004 | 0.0061 | 0.0022 | 0.0578 | 1 |
| 4 | Belarus | 0.6389 | 1.6081 | 0.0054 | 0.0054 | 0.0556 | 1 |
| 5 | Belgium | 0.5913 | 1.4518 | 0.0115 | 0.0005 | 0.0501 | 3 |
| 6 | Brazil | 0.4642 | 1.6686 | 0.0060 | 0.0088 | 0.0512 | 1 |
| 7 | Canada | 0.5949 | 1.5198 | 0.0098 | 0.0024 | 0.0507 | 3 |
| 8 | Chile | 0.4217 | 1.4200 | 0.0106 | 0.0039 | 0.0532 | 3 |
| 9 | China | 0.3623 | 1.5113 | 0.0139 | 0.0139 | 0.0440 | 2 |
| 10 | Colombia | 0.5754 | 1.3792 | 0.0123 | 0.0005 | 0.0594 | 3 |
| 11 | Czech Republic | 0.5676 | 1.3824 | 0.0135 | 0.0002 | 0.0579 | 3 |
| 12 | Denmark | 0.4225 | 1.4223 | 0.0110 | 0.0057 | 0.0547 | 3 |
| 13 | Dominican Republic | 0.6034 | 1.3572 | 0.0089 | 0.0000 | 0.0582 | 3 |
| 14 | Ecuador | 0.6140 | 1.4507 | 0.0079 | 0.0016 | 0.0501 | 3 |
| 15 | Egypt | 0.4160 | 1.3666 | 0.0154 | 0.0006 | 0.0589 | 3 |
| 16 | Finland | 0.4179 | 1.3641 | 0.0128 | 0.0032 | 0.0579 | 3 |
| 17 | France | 0.6182 | 1.5399 | 0.0080 | 0.0013 | 0.0465 | 1 |
| 18 | Germany | 0.6058 | 1.5082 | 0.0092 | 0.0045 | 0.0447 | 1 |
| 19 | India | 0.4606 | 1.6236 | 0.0071 | 0.0032 | 0.0535 | 1 |
| 20 | Indonesia | 0.5935 | 1.3815 | 0.0100 | 0.0012 | 0.0561 | 3 |
| 21 | Iran | 0.5900 | 1.5097 | 0.0103 | 0.0035 | 0.0475 | 1 |
| 22 | Ireland | 0.6110 | 1.5126 | 0.0083 | 0.0037 | 0.0538 | 3 |
| 23 | Israel | 0.5877 | 1.4351 | 0.0106 | 0.0015 | 0.0542 | 3 |
| 24 | Italy | 0.6146 | 1.6088 | 0.0087 | 0.0011 | 0.0479 | 1 |
| 25 | Japan | 0.5892 | 1.4282 | 0.0104 | 0.0012 | 0.0542 | 3 |
| 26 | Korea | 0.8114 | 1.4576 | 0.0138 | 0.0221 | 0.0485 | 4 |
| 27 | Malaysia | 0.4385 | 1.4242 | 0.0122 | 0.0058 | 0.0570 | 3 |
| 28 | Mexico | 0.6508 | 1.6937 | 0.0067 | 0.0141 | 0.0539 | 1 |
| 29 | Netherlands | 0.6252 | 1.4704 | 0.0068 | 0.0000 | 0.0530 | 3 |
| 30 | Norway | 0.5685 | 1.3809 | 0.0125 | 0.0000 | 0.0580 | 3 |
| 31 | Pakistan | 0.4336 | 1.4382 | 0.0095 | 0.0018 | 0.0539 | 3 |
| 32 | Panama | 0.6167 | 1.3813 | 0.0092 | 0.0000 | 0.0597 | 3 |
| 33 | Peru | 0.4285 | 1.5118 | 0.0098 | 0.0051 | 0.0508 | 3 |
| 34 | Philippines | 0.5984 | 1.3850 | 0.0102 | 0.0000 | 0.0583 | 3 |
| 35 | Poland | 0.5922 | 1.4366 | 0.0101 | 0.0021 | 0.0548 | 3 |
| 36 | Portugal | 0.6082 | 1.4464 | 0.0085 | 0.0000 | 0.0537 | 3 |
| 37 | Qatar | 0.6440 | 1.5227 | 0.0057 | 0.0038 | 0.0551 | 1 |
| 38 | Romania | 0.5940 | 1.3970 | 0.0099 | 0.0027 | 0.0543 | 3 |
| 39 | Russian Federation | 0.6618 | 1.6205 | 0.0035 | 0.0005 | 0.0489 | 1 |
| 40 | Saudi Arabia | 0.4432 | 1.4619 | 0.0075 | 0.0008 | 0.0534 | 3 |
| 41 | Serbia | 0.6218 | 1.3999 | 0.0072 | 0.0003 | 0.0579 | 3 |
| 42 | Singapore | 0.6632 | 1.5509 | 0.0028 | 0.0028 | 0.0543 | 1 |
| 43 | South Africa | 0.4348 | 1.4128 | 0.0113 | 0.0022 | 0.0601 | 3 |
| 44 | Spain | 0.6104 | 1.5719 | 0.0085 | 0.0020 | 0.0462 | 1 |
| 45 | Sweden | 0.6028 | 1.4258 | 0.0097 | 0.0000 | 0.0539 | 3 |
| 46 | Switzerland | 0.2720 | 1.5551 | 0.0103 | 0.0098 | 0.0507 | 2 |
| 47 | Turkey | 0.6427 | 1.6682 | 0.0052 | 0.0010 | 0.0519 | 1 |
| 48 | Ukraine | 0.6649 | 1.6096 | 0.0033 | 0.0030 | 0.0574 | 1 |
| 49 | United Arab Emirates | 0.4399 | 1.4730 | 0.0098 | 0.0021 | 0.0551 | 3 |
| 50 | United Kingdom | 0.6256 | 1.5004 | 0.0068 | 0.0000 | 0.0464 | 1 |
| 51 | United States | 0.6304 | 1.6520 | 0.0063 | 0.0001 | 0.0436 | 1 |

